# ACE-2-like Enzymatic Activity in COVID-19 Convalescents with Persistent Pulmonary Symptoms Associated with Immunoglobulin

**DOI:** 10.1101/2025.02.12.25322167

**Authors:** Yufeng Song, Frances Mehl, Lyndsey M. Muehling, Glenda Canderan, Kyle Enfield, Jie Sun, Michael T. Yin, Sarah J. Ratcliffe, Jeffrey M. Wilson, Alexandra Kadl, Judith A. Woodfolk, Steven L. Zeichner

## Abstract

Many difficult to understand clinical features characterize COVID-19 and Post-Acute Sequelae of COVID-19 (PASC or Long COVID, LC). These can include blood pressure instability, hyperinflammation, coagulopathies, and neuropsychiatric complaints. The pathogenesis of these features remains unclear. The SARS-CoV-2 Spike protein Receptor Binding Domain (RBD) binds Angiotensin Converting Enzyme 2 (ACE2) on the surface of host cells to initiate infection. We hypothesized that some people convalescing from COVID-19 may produce anti-RBD antibodies that resemble ACE2 sufficiently to have ACE2-like catalytic activity, that is they are ACE2-like proteolytic abzymes that may help mediate the pathogenesis of COVID-19 and LC. In previous work, we showed that some people with acute COVID-19 had immunoglobulin-associated ACE2-like proteolytic activity, suggesting that some people with COVID-19 indeed produced ACE2-like abzymes. However, it remained unknown whether ACE2-like abzymes were seen only in acute COVID-19 or whether ACE2-like abzymes could also be identified in people convalescing from COVID-19. Here we show that some people convalescing from COVID-19 attending a clinic for people with persistent pulmonary symptoms also have ACE2-like abzymes and that the presence of ACE2-like catalytic activity correlates with alterations in blood pressure in an exercise test.

## Introduction

The COVID-19 pandemic has had a devastating effect on world health. An estimated 7M have died so far ^1^, even with the availability of effective vaccines and antivirals. Beyond COVID-19’s acute effect, its long-term effects have had grave effects on individuals and society. Among the most challenging secondary consequences of COVID-19 are people affected by Post-Acute Sequelae of COVID-19 (PASC, or Long COVID, LC), reviewed in ^2–4^, which may affect as many as 7% of people with COVID-19 ^5^. Many studies have described the epidemiology and clinical manifestations of LC, and associated LC with various clinical findings, but no clear fundamental, causal mechanisms have been established ^6^. LC symptoms can include post-exertional malaise, fatigue, “brain fog” and other neuropsychiatric symptoms, dizziness, GI complaints, cardiovascular complaints like palpitations, libido changes, alterations in smell or taste, cough, chest pain, and abnormal movements ^7^. Many people with LC exhibit abnormal clinical features, including blood pressure dysregulation, coagulopathies, and inflammation. ^8, 9^. People with LC have been found to have distinct immune profiles compared to matched subjects without LC, including altered populations of lymphoid and myeloid cells, altered levels of immune mediators and hormones and increased humoral immune responses against SARS-CoV-2 ^10^. Proteomic studies have shown that people with LC have evidence of persistent activation of the complement (C’) system with increased concentrations of complement proteolysis products and proteolysis by thrombin ^11^, but these findings have not identified a proximate cause underlying the disease. Since no proximate cause of LC has been identified, no specific therapies are available, and people with LC continue to suffer and are treated only with non-specific therapies directed at one or another symptom.

We noted that proteolytic regulatory cascades control many of the physiologic pathways dysregulated in COVID-19 and LC, such as blood pressure regulation (angiotensin), coagulation (thrombin), and inflammation (C’, kallikrein-kinin) ^12–14^. Some of the problematic clinical disorders related to COVID-19 do not become apparent until a week or more after infection, so their underlying pathogenesis is likely due to secondary or reactive processes and not due to primary cytotoxicity from acute viral replication. Additional data suggests that some of the COVID-19-associated inflammatory processes may involve antibodies against SARS-CoV-2 spike protein, S. For example, a small number of individuals who received SARS-CoV-2 vaccines and had no evidence of SARS-CoV-2 infection exhibit disordered inflammatory responses, blood pressure dysregulation or coagulopathy^15^, and a small number of infants born to mothers with COVID-19 have been described as having multi-system inflammatory syndrome of children (MIS-C)^16^.

Since many of COVID-19 and LC’s puzzling features appear to involve physiologic regulatory cascades controlled by proteolysis, we considered how infection with SARS-CoV-2 might yield pathologies involving proteolytic regulatory cascades. We noted that angiotensin converting enzyme 2 (ACE2) is SARS-CoV-2’s host cell receptor ^17^. The Receptor Binding Domain (RBD) of the SARS-CoV-2 Spike protein (S), the viral envelope protein responsible for mediating virus binding and entry, has interactions with ACE2 that include the active site^18–22^. ACE2 exists both free in the circulation and in the membranes of cells of several tissues. While angiotensin converting enzyme (ACE) cleaves angiotensin I to yield angiotensin II, a vasoconstrictor that increases blood pressure ^23^, ACE2 acts in a counter-regulatory fashion to ACE, cleaving angiotensin II to yield angiotensin 1-7, a vasodilator that lowers blood pressure ^24^. We recalled the literature concerning catalytic antibodies or “abzymes” ^25, 26^. When abzymes were first identified, researchers created abzymes by making anti-idiotypic antibodies against antibodies that themselves had been produced against the active site of a particular enzyme of interest^27, 28^. Some of the anti-idiotypic antibodies resembled the enzyme’s active site sufficiently for the antibody to have catalytic activity like that of the original enzyme^29, 30^. However, some abzymes exhibited substrate specificities that differed notably from those of the original enzyme’s ^28, 29, 31^. The abzymes exhibited substrate promiscuity compared to the original enzyme and had catalytic activity that was lower than the activity typically seen with more conventional enzymes (*k*cat/*Km* values ∼10^2^ - 10^4^ s^-1^•M s^-1^ ^32^ vs. *k*cat/*Km* values of ∼10^5^ s^-1^•M s^-1^)^33^. The poorer activity, in addition to the problems with substrate specificity spread, led to decreased interest in abzymes as a new biotechnology.

We hypothesized that some people with COVID-19 may develop antibodies with catalytic activity because the RBD, since it binds ACE2, should have a partial negative image of ACE2, so some anti-RBD antibodies may have enough similarity to ACE2 to have proteolytic activity similar to ACE2, or perhaps other proteolytic enzymes due to substrate specificity spread seen with abzymes. ACE2 can cleave many peptides with important physiologic activity beyond angiotensin II^34^.

There are examples of people who produce abzymes with potentially clinically significant effects, for example DNA ^35^, immunoglobulin ^36^, vasoactive intestinal peptide (VIP)^37^, myelin basic protein (MBP) in persons with multiple sclerosis^38^, and factors in the coagulation cascade ^39–41^. Some people with HIV have been shown to produce abzymes that cleave HIV Env, so there exist examples of cases where viral infection induces production of an abzyme ^42^.

In previous work ^43^, we showed that some people with acute COVID-19 produced antibodies with ACE2-like catalytic activity. However, the question remained whether antibodies with ACE2-like catalytic activity were present in people convalescing from COVID-19 and, if so, whether the presence of ACE2-like catalytic activity correlated with any plausible clinical findings. Here we show results obtained from 20 people convalescing from COVID-19 who attended a clinic at the University of Virginia for people convalescing from COVID-19 with persistent pulmonary symptoms. Prior work identified persistent inflammation and immune dysregulation within these individuals, including auto-reactive antibodies ^44^. We found that six of those volunteers had ACE2-like antibody-associated catalytic activity in their plasma, and that the presence of ACE2-like antibody-associated catalytic activity correlated with lower blood pressure after a six-minute walk test, indicating that ACE2-like abzyme activity occurs in people with both acute COVID-19 and people convalescing from COVID-19 and can have associations with physiologic consequences.

## Methods

### Clinical Cohort

We studied a subset of individuals from a previously published cohort of people convalescing from COVID-19 at the University of Virginia^44^ who attended a post-COVID pulmonary clinic for follow-up care (COVID Recovery Cohort (COVID-RC)). All research subjects had confirmed prior COVID-19 infection. Samples and other physiologic monitoring were obtained after informed consent. The University of Virginia Institutional Review Board for Health Sciences Research (FWA #00006183) approved enrollment of all subjects and collection of specimens and related metadata (HSR #13166) and approval was obtained to work on the specimens (HSR #HSR200362). Twenty research subjects, spanning a broad range of anti-Spike IgG levels (2.1-314.6 *μ*g/mL), with available plasma biospecimen, were selected for abzyme studies. The characteristics of the research volunteers selected for abzyme studies are listed in Table 1. Research subjects had been hospitalized during acute infection (70%), with 30% requiring mechanical ventilation. The research volunteers initially contracted COVID-19 from March 2020-March 2022, with the majority becoming infected prior to the circulation of major viral variants (70%). SARS-CoV-2 anti-spike and anti-capsid IgG was measured by ImmunoCAP assay^43, 45^. Approximately half of the selected subjects (55%) had received at least one SARS-CoV-2 vaccine dose at the time of sample collection. While there was a statistically significant association between receipt of at least one dose of vaccine and anti-spike IgG, there was no significant association between vaccine receipt and the presence of ACE-2-like catalytic antibodies and no statistically significant association between anti-spike IgG level and the presence of ACE-2-like catalytic antibodies We purchased EDTA-anticoagulated healthy normal donor plasma from Valley Biomedical (http://www.valleybiomedical.com/; Pooled Human Plasma, Cat. No. HP1051PK2, Lot No. 21M2548). Valley Biomedical collected plasma for this lot on the 12th and 13th of December 2018 (personal communication with Valley Biomedical technical support), about a year before the first cases of COVID-19.

**Table 1.** Demographic and clinical characteristics of the research volunteers included in the study.

| Subject Characteristics (n=20) |  |
| --- | --- |
| Age(yrs) | 54+/-19 |
| Sex | Male (55%), Female (45%) |
| Race | White (85%), Black (10%), Other (5%) |
| Smoking history | Former (30%), Never (70%) |
| Days since start of illness | 201 +/-129 |
| Hospitalized for Covid | Yes (70%), No (30%) |
| History of hypertension | Yes (35%), No (65%) |
| Cardiovascular conditions | Bradycardia (10%), Heart murmur (5%), Aortic stenosis (5%), None (85%) |
| Pulmonary conditions | Asthma (35%), Obstructive sleep apnea (20%), COPD (5%), IPF (5%), Sarcoidosis (5%), None (50%) |
| Autonomic conditions | Yes (5%), No (95%) |
| BMI | 32.57+/-7.49 |
| Hyperlipidemia | Yes (25%), No (75%) |
| Diabetes | Type 2 (25%), Pre (5%), Steroid-induced (5%), None (65%) |
| GERD | Yes (35%), No (65%) |
| Other Conditions | Hx Cancer (15%), Osteoarthritis (15%), Hypothyroidism (10%), CKD (5%), Gout (5%), HIV (5%), NASH (5%), RA (5%), None (55%) |

### Clinical and Other Data

Clinical data collected from the volunteers in the cohort included a symptom inventory. A six-minute walk test was given to 18 of the subjects included in this study, in which heart rate, systolic and diastolic blood pressures were recorded before and after the test walk. Data on plasma cytokines TNF-α and IL-6 was obtained for 12 volunteers, as previously described^44^.To explore whether inflammatory cytokine markers might correlate with the presence of ACE2-like catalytic activity, we examined TNF-α and IL-6 levels for some of the subjects, but found no statistically significant correlations. Table 2 summarizes this data.

**Table 2.** Symptoms, cytokine assays, and physiologic measurements conducted on the subjects in the study. For the data listed under the six-Minute Walk Test heading, Pre-HR is the heart rate before the walk, Post-HR is the heartrate after the walk, Change HR is (PostHR – PreHR), Pre SBP is the systolic blood pressure before the walk, Post SBP is the systolic blood pressure after the walk, Change SBP is (Pre SBP – Post SBP), Pre DBP is the diastolic blood pressure before the walk, Post DBP is the diastolic blood pressure after the walk, Change DBP is (Pre DBP – Post DBP).

| **Symptoms** |  |
| --- | --- |
| *Pulmonary Symptoms* | *People with Symptom* |
| Cough | 40% |
| Breathing Difficulty | 10% |
| Dyspnea on exertion | 80% |
| *Other Symptoms* |  |
| Headache | 10% |
| Weakness | 15% |
| Fatigue | 60% |
| Dizzy/Lightheaded/Syncope | 20% |
| Brain fog/Memory Problems | 35% |
| Sleep Disruption | 45% |
| Pain | 25% |
| Numbness | 15% |
| **Inflammatory Cytokines (n=12)** |  |
| TNF-α (pg/mL) | 58 +/-52 |
| IL-6 (pg/mL) | 3.2 +/-3.2 |
| **Six-Minute Walk Test (n=18)** |  |
| Pre-HR (beats/minute) | 88 +/-12 |
| Post-HR (beats/minute) | 111 +/-19 |
| Change HR (beats/minute) | 23 +/-13 |
| Pre SBP (mmHg) | 137 +/-17 |
| Post SBP (mmHg) | 145 +/-23 |
| Change SBP (mmHg) | 8 +/-16 |
| Pre DBP (mmHg) | 80 +/-8 |
| Post DBP (mmHg) | 82 +/-12 |
| Change DBP (mmHg) | 2 +/-9 |

### Angiotensin II Converting Enzyme (ACE2)-like Abzyme Activity Detection

Abzyme catalytic activities in plasma samples were measured using an Angiotensin II Converting Enzyme (ACE2) Activity Assay Kit, Fluorometric (Abcam, Cat. #ab273297) providing a synthetic ACE2 peptide substrate labeled with a fluor and a quencher with a lower limit of detection for an unmodified assay of 400 µU as described ^43^. Recombinant human ACE2 (ACE2) as a positive control is also included in the assay kit. ACE2 inhibitor provided in the assay kit was also used for additional controls. Briefly, 50*μ*l plasma or diluted plasma (in PBS without Ca^2+^ or Mg^2+^, and with 4mM EDTA added) was added to wells in a 96-well black flat bottom assay plate (Corning, Cat. #3603) and incubated at room temperature (RT), in the dark for 15 min. Then, 50*μ*l pre-diluted substrate was added into the wells of the plate, mixing the substrate with the plasma sample. The plate was immediately placed into a SpectraMax® M5 multi-model microplate reader (Molecular Devices). After 3 minutes incubation, the Relative Fluorescent Unit (RFU, Ex/Em = 320/420 nm) values were measured in kinetic mode every 20 min for a total of 16 hours at 37℃. We performed assays on normal human plasma spiked with serial dilutions of rACE2 to determine the lower limits of detection for the assay. We then defined as a “positive” samples that had RFU slopes (between 5 and 120 minutes) that were greater than or equal to 2-fold the slope of the lower limit of detection of serial dilution assays using rACE2 spiked into normal human plasma. One of the samples, number 10, had an anomalous result where the activity initially increased, then decreased. We repeated the assays for this sample and obtained equivalent results. We do not have an explanation for this anomalous result, but did note that the sample well was not as clear as the other samples, so it is possible that there was some sort of contaminating material or activity, but we were not able to determine what it was. Nevertheless, in the interest of completeness, we include the data from this sample in our dataset.

We conducted negative control experiments using the ACE2 inhibitor from the assay kit for additional controls.

### Human Immunoglobulin Depletion and Detection

We depleted immunoglobulin from the plasma samples as previously described in detail ^43^. Plasma samples were 1∶8 diluted in PBS with 4 mM EDTA and centrifuged at 12,000 × g at 4 ℃ for 15 min followed by passing through 0.45 *μ*m syringe filter (FisherScientific, Cat. #97204). We loaded 500 *μ*L of the filtered samples onto ultrafiltration columns with 100kDa cut-off ultrafiltration membrane (Pierce™ Cat. # 88503) and centrifuged the columns at 12,000 × g at 4 ℃ until > 400 *μ*L of filtrate was produced. The flow-through fractions were then incubated with 30 *μ*L pre-washed Protein A/G Magnetic Beads (Life Technologies, Cat. # 88803, 10 mg/mL) at room temperature (RT) for 1 hour, and repeated a second time to further deplete the antibodies. Human IgG was measured using a Human IgG ELISA Kit (Abcam, Cat. #ab100547) according to the manufacturer’s protocol as described ^43^. Briefly, 100 *μ*l samples, as well as standards, were added into each well of multi-microwell strips and incubated at room temperature for 2.5 hours. Biotin-labeled IgG detection antibody was then added to each well after wash and incubated at room temperature for 1 hour. Then, HRP-streptavidin was added at room temperature for 1 hour. After wash, OD values were developed by adding TMB One-Step Substrate and Stop Solution subsequently. OD values were read at 450 nm using an accuSkan FC micro-well plate reader (ThermoFisher). IgG concentrations were calculated based on standard curves generated with the same batch of assay strips. Our depletion protocol removes >99.99% of human IgG and IgM from the plasma samples^43^.

### ACE2 Substrate Peptide Cleavage Competition Inhibition by SARS-CoV-2 Spike RBD Peptide Pools

As described previously ^43^ we synthesized tiled SARS-CoV-2 spike protein RBD (receptor binding domain) peptides (SB-PEPTIDE, SmartBioscience SAS, France) from residues Arg319 to Phe541, including receptor binding motif (RBM) from Ser438 to Gln506, based on a published analysis on viral-host interaction ^21^. We added 10*μ*g per peptide to selected plasma samples, diluted 1∶8 dilution with PBS with 4 mM EDTA. Recombinant human ACE2 protein, supplied in the kit served as a positive control. Sample-peptide pool mixtures were incubated at RT for 15 min in dark before adding the substrate. Samples incubated with ddH_2_O and DMSO (1∶1 mixture), equivalent to the concentrations of the peptide pool reconstitution served as negative controls. Relative Fluorescent Units were measured as described above, and data was analyzed as described below.

### Data Analysis

As previously described ^43^, RFU values observed over 120 min were used to evaluate ACE2 substrate cleavage activities in those plasma samples. Corrected value of each sample was calculated by subtracting baseline RFU value from the value at each subsequent time point and the sum of baseline-corrected value over 120 min was also calculated using R. The change in RFU over the first 120 minutes of the assay was calculated using linear regression and was used as a measure of ACE2-like activity. Correlations between ACE2 like activity and clinical data were assessed using Pearson’s product-moment correlation for numerical variables and Kruskal-Wallis rank/sum test for categorical variables. Data analysis and visualization was performed with R (version 2022-06-23) using the Rstudio user interface with packages including ggplot2, tidyverse, readr, drc, ggpubr, ggh4x, stringr, and patchwork.

## Results

To determine whether individuals convalescing after COVID-19 produced antibodies with ACE2-like catalytic activity, similar to those we identified in people with acute COVID-19^43^, we studied samples available from 20 individual from an existing cohort available at the University of Virginia for which plasma samples and clinical testing was available. This cohort, already described^44^, enrolled people convalescing from COVID-19, the majority of whom reported persistent pulmonary symptoms. The demographic characteristics of the cohort are described in Table 1.

We conducted ACE2 activity assays on 20 plasma samples from these research subjects. We found that 6 of them had ACE2-like catalytic activity greater than or equal to 2-fold above the lower limits of detection for ACE2 as determined by serial dilution experiments using rACE2, our criteria for positive ACE2-like activity. An additional 5 subjects had borderline positive ACE2-like activity, with levels greater than or equal to the lower limits of detection of the rACE2, but less than the 2-fold greater than the lower limit of detection cut-off (FIGs 1 and 2).

**FIG 1.**
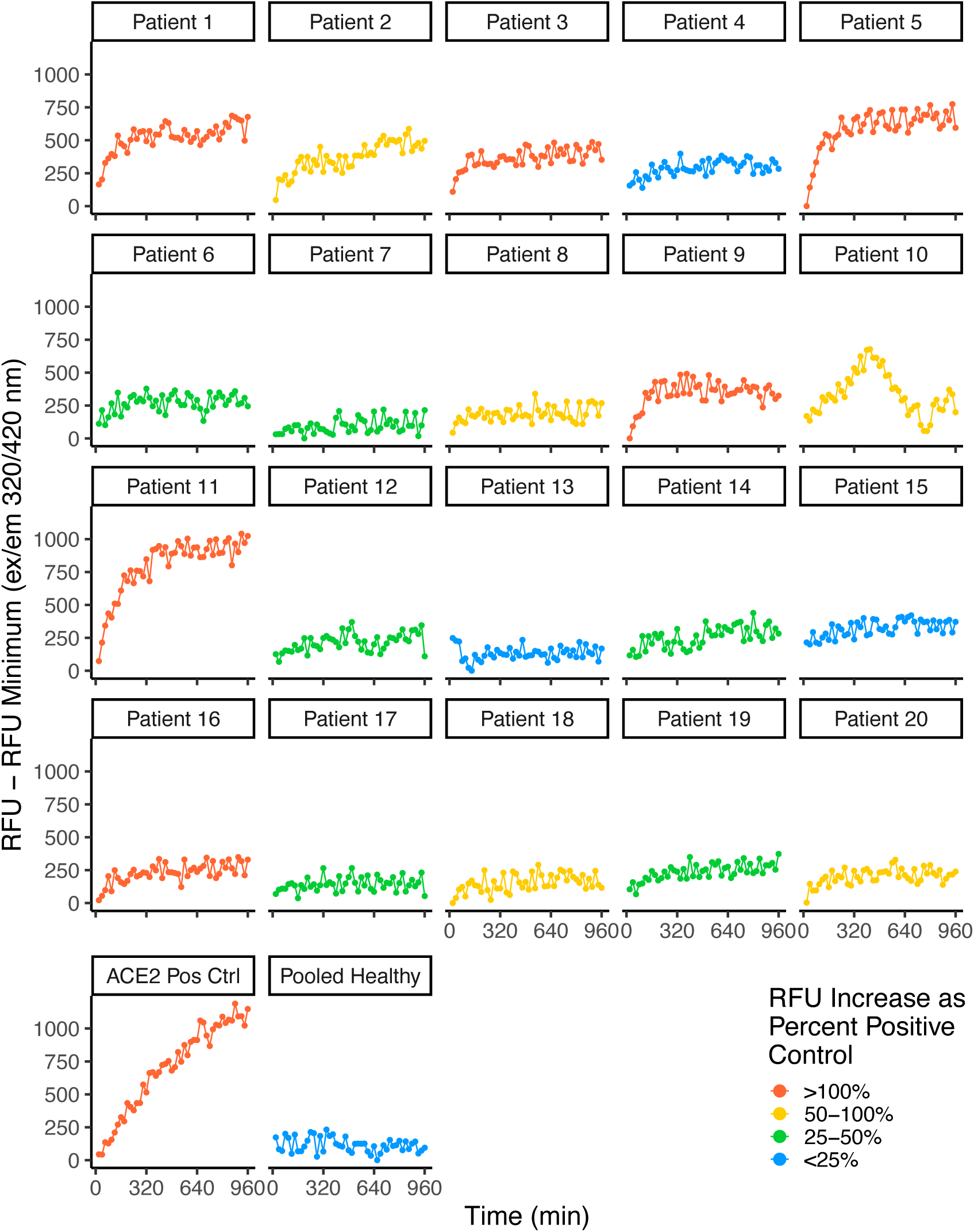
ACE-2-Like Activity in Convalescent Patient Plasma. Assays for ACE2-like enzymatic activity conducted on EDTA-anti-coagulated plasma from 20 COVID-19 convalescent volunteers with persistent pulmonary symptoms, conducted with a fluor-quench tagged peptide substrate, as described in ^43^. We determined the assay slopes over 5-120 minutes and then called as a positive any sample with a 5–120-minute slope greater than twice the slope of the lower limit of detection of the positive control (recombinant ACE2).

**FIG 2.**
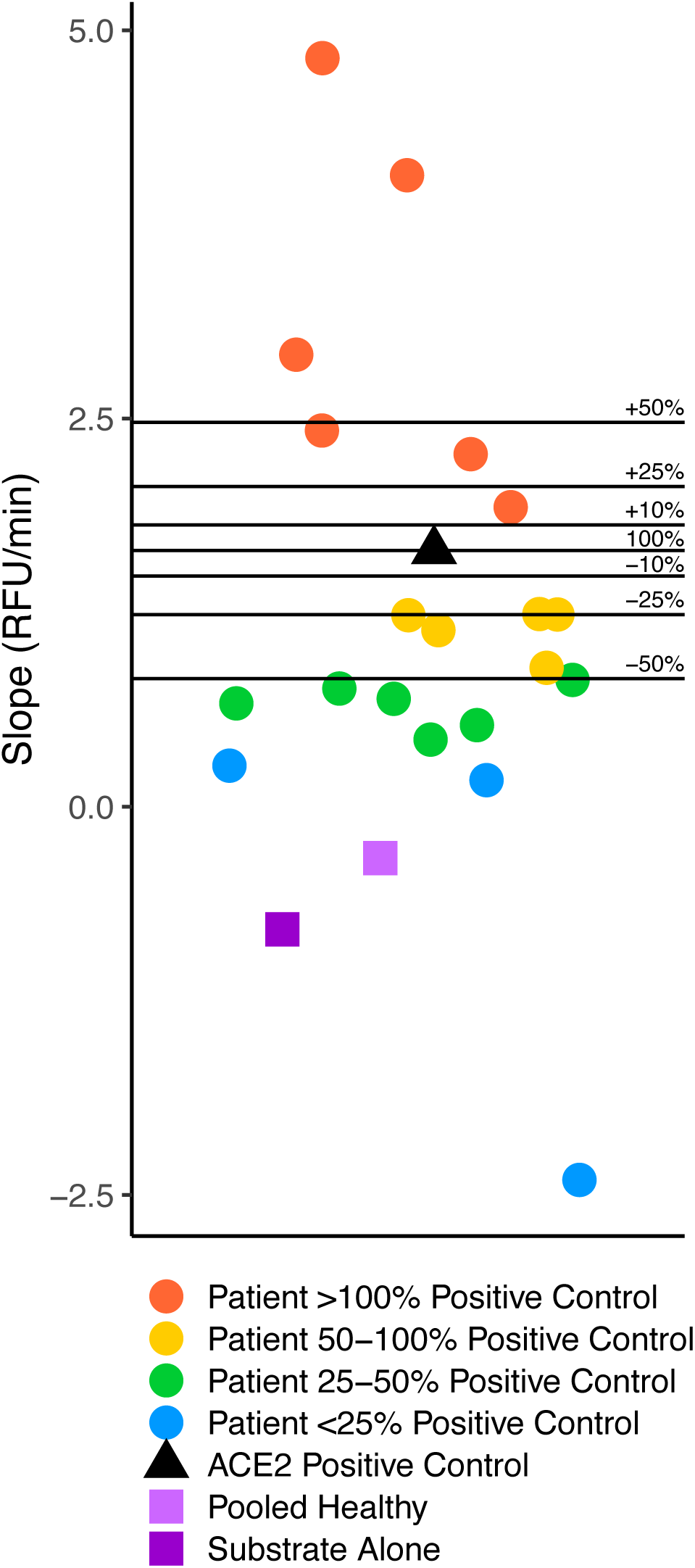
Distribution of ACE-2-like activity in the Convalescent Research Subjects. Distribution of the 5-120 minute ACE2-like enzymatic activity slopes from Fig 1, compared to the recombinant human rACE2 positive control. We found that 6 out of the 20 plasma samples had slopes that we considered positive, with slopes greater than that observed for the rACE2 positive control at twice the lower limit of detection. An additional 5 subjects had ACE2-like catalytic activity with 5–120-minute slopes (RFU/min) greater than or equal to the slope observed with 0.06 ng rACE2 at the lower limit of detection.

To determine whether the ACE2-like activity was associated with immunoglobulin in these samples, we conducted an immunoglobulin depletion procedure, that included depleting molecules >100 KDa using a centrifugal ultrafiltration column followed by a protein A/G absorption step. The procedure yielded samples that were depleted in immunoglobulin by >99.9% (Table 3).

**Table 3.** Immunoglobulin depletion following high molecular weight depletion and protein A/G bead absorption. Assays for the IgG concentration were conducted in triplicate.

| Patient | Plasma IgG Concentration<br>Mean +/-SD |  | IgG<br>Depletion<br>Efficiency |
| --- | --- | --- | --- |
|  | Pre-IgG<br>Depletion<br>(mg/ml) | Post-IgG Depletion<br>(ng/ml) |  |
| #1 | 1.62 +/-0.01 | 0.19 +/-<0.001 | >99.99% |
| #3 | 0.40 +/-0.01 | 0.19 +/-<0.001 | >99.99% |
| #5 | 1.82 +/-0.02 | 0.19 +/-<0.001 | >99.99% |
| #9 | 0.82 +/-0.05 | 0.19 +/-<0.001 | >99.99% |
| #11 | 0.53 +/-0.01 | 0.19 +/-<0.001 | >99.99% |
| #16 | 0.45 +/-0.08 | 0.19 +/-0.003 | >99.99% |
| #2 | 1.03 +/-0.01 | 0.19 +/-0.000 | >99.99% |
| #20 | 0.63 +/-0.04 | 0.19 +/-0.001 | >99.99% |
| #14 | 1.27 +/-0.02 | 0.18 +/-0.001 | >99.99% |
| #13 | 0.60 +/-0.01 | 0.18 +/-0.001 | >99.99% |
| HDP | 2.86 +/-1.72 | 0.03 +/-0.004 | >99.99% |

We conducted the ACE2-like activity assays on the immunoglobulin-depleted plasma samples from the subjects who were found to be positive for ACE2-like activity. The immunoglobulin-depleted samples had no ACE2-like activity, supporting the hypothesis that the ACE2-like activity is antibody-associated (FIG 3).

**FIG 3.**
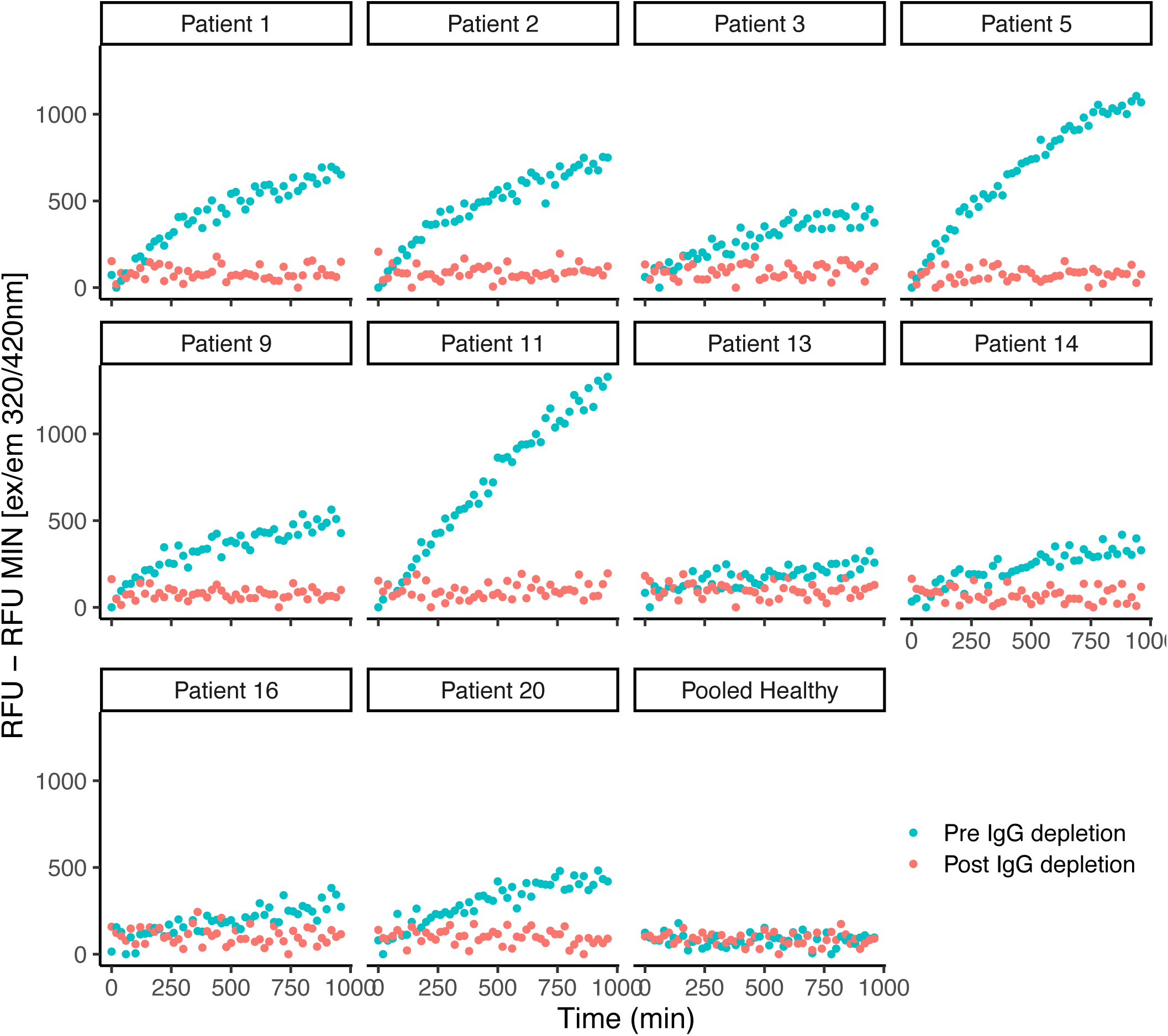
ACE2-like Activity in Samples Depleted of Immunoglobulin. Assays for ACE2-like enzymatic activity conducted on EDTA-anti-coagulated plasma of the 6 out of 20 COVID-19 convalescent research volunteers with persistent pulmonary symptoms positive for ACE2-like activity in FIGs 1 and 2, conducted with a fluor-quench tagged peptide substrate, with immunoglobulins depleted as described in the Methods and ^43^.

To further confirm the specificity of the ACE2-like activity, we conducted assays on the samples in the presence of tiled overlapping peptides that included the RBD site in the spike protein ^43^. Competition with using the overlapping spike RBD region peptides strongly inhibited the ACE2-like cleavage activity present in the patient plasma samples, further confirming that the ACE2-like activity was specific for an RBD-interacting activity of the plasma antibodies. There was a modest decrease when the competing peptide diluent was added alone, probably because addition of the diluent increased the volume of the material assayed in the wells (FIG 4).

**FIG 4.**
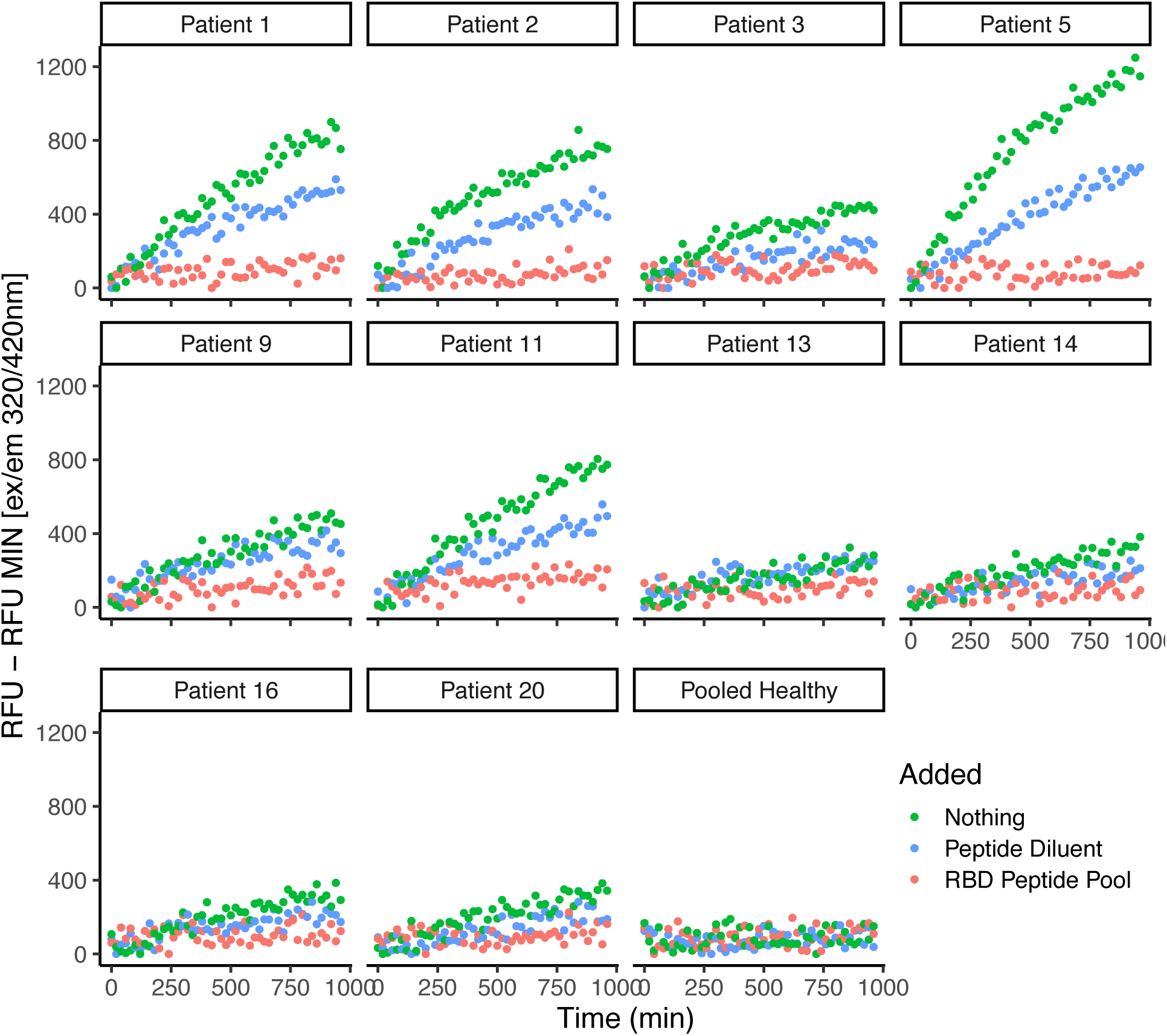
Inhibition of ACE2-like catalytic activity in plasma from COVID-19 convalescent research subjects by pooled tiled RBD peptides. Assays for ACE2-like enzymatic activity conducted on EDTA-anti-coagulated plasma from 20 COVID-19 convalescent individuals with persistent pulmonary symptoms, conducted with a fluor-quench tagged peptide substrate, with ACE2 activity competed using tiled overlapping RBD peptides as described in ^43^.

The cohort studied here consisted of individuals with persistent pulmonary symptoms (complaints of cough, difficulty breathing, or dyspnea on exertion) not subjects enrolled specifically because they had symptoms that could be considered characteristic for LC. Nevertheless, several individuals in this cohort had a variety of symptoms beyond the strictly pulmonary symptoms. Table 2 lists the range of symptoms reported in this cohort.

If SARS-CoV-2 infection elicits the production of abzymes, it would be important to know if those abzymes were associated with any clinical features. Here, we focused on catalytic antibodies with ACE2-like activity, so for this study our interests centered on whether there would be a correlation in the research volunteers between ACE2-like catalytic activity and a physiologic variable that could be related to dysregulated ACE2-like activity. ACE2 catalyzes the cleavage of the vasoconstrictor angiotensin II to the counterregulatory vasodilator angiotensin 1-7. A dysregulation of angiotensin 1-7 production might be expected to prevent the maintenance of ideal vascular tone. So, if an individual has an ACE2-like catalytic antibody in the circulation, we hypothesized that they may experience problems with blood pressure regulation, particularly after an event that demanded a physiologic response to maintain blood pressure homeostasis, like an exercise challenge. Thus, we asked whether research volunteers with higher ACE2-like abzyme activities were less likely to maintain their blood pressures during a 6-minute walk test. Analysis of data from 18 volunteers revealed a significant correlation between the change in systolic blood pressure after 6-minute walk test and ACE2-like activity, with subjects showing higher ACE2-like activity experiencing greater decreases in systolic blood pressure (r=-0.54, p=0.021). Non-statistically significant trends associated ACE-2 like catalytic activity with an increase in heart rate after the six-minute walk test and the absolute systolic blood pressure after six-minute walk test (FIG 5).

**FIG 5.**
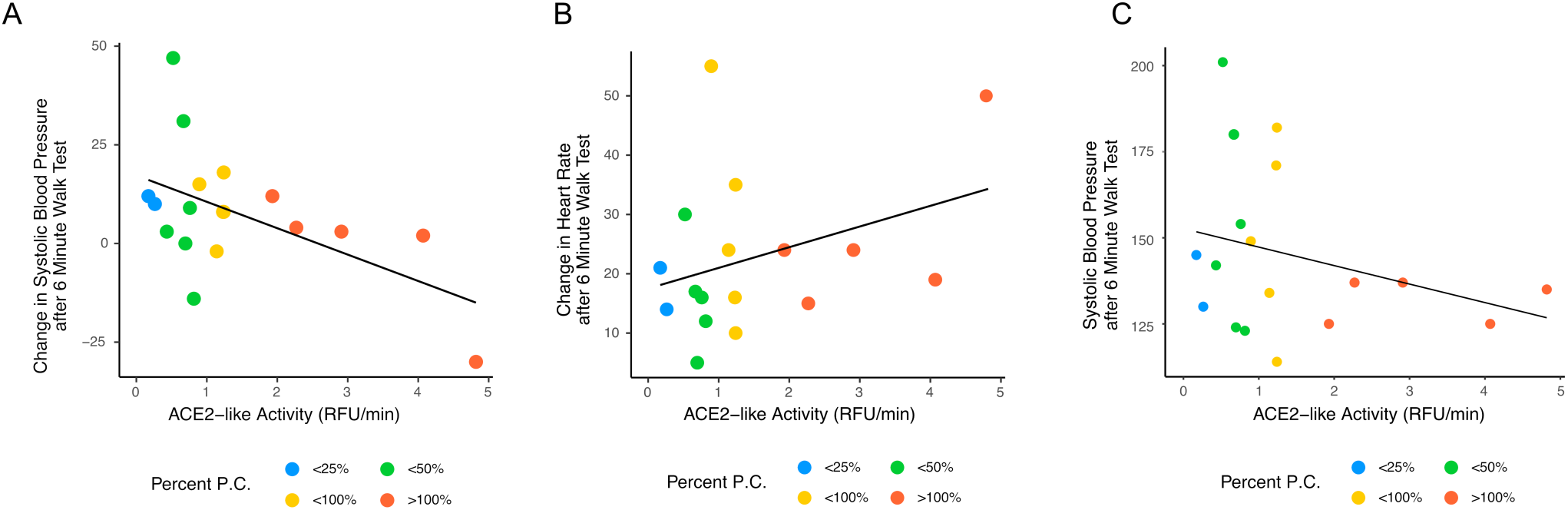
Correlation between cardiovascular six-minute walk stress test variables and ACE2-like catalytic activity. A. Change in systolic blood pressure vs. ACE2-like catalytic activity. B. Change in heart rate vs. ACE2-like catalytic activity. C. Systolic blood pressure vs. ACE2-like catalytic activity. The change in systolic BP had a correlation of -0.54, p = 0.021. The changes in heart rate (correlation -0.29, p = 0.241) and absolute systolic blood pressure after the 6-minute walk (correlation 0.34, p = 0.178) were not significant (Pearson’s product moment correlation).

## Discussion

Our prior work ^43^ showed that some people with acute COVID-19 produced antibodies that had ACE2 catalytic activity (ACE2-like abzymes) and that abzyme activity correlated with RBD binding activity. However, that study involved only individuals with acute COVID-19, and did not address whether the ACE2-like abzyme activity persisted after resolution of the acute disease into the convalescent period. In this study, we show that some people convalescing from COVID-19 also have antibodies with ACE2-like abzyme activity, and that the presence of ACE2-like abzyme activity correlates with a physiologically plausible clinical finding, namely decrease in blood pressure after six-minutes of walking.

The finding that people convalescing from COVID-19 can have circulating ACE2-like abzymes that correlate with a clinical finding supports the hypothesis that post-COVID-19 circulating abzymes could be involved in other aspects of disorders observed in people convalescing from COVID-19. These could conceivably include some of the clinical features of LC, since many of the clinical problems observed in people with LC include features that can be mediated by proteolytic regulatory cascades, although considerable additional work on larger cohorts with different clinical manifestations of LC, and additional assays for other antibody-associated catalytic activity would be needed to establish such associations. However, the hypothesis is highly testable, able to inform clear, follow-on studies, such as studies to determine if people convalescing from COVID-19, with LC, have circulating abzymes that can affect additional proteolytic regulatory cascades capable of mediating one or another aspect of LC pathogenesis. If pathophysiologically active SARS-CoV-2-induced abzymes are found, they could offer a plausible target for the development of new therapies, which could be useful, since there are no currently available therapies directed at a proximate cause of LC.

If there are some antibodies produced in people with COVID-19 that have ACE2-like catalytic activity, it might be reasonable to hypothesize that some people with COVID-19 might also make antibodies against the RBD that could inhibit ACE2 activity. Indeed, there has been a report of such antibodies ^46^. Findings like these further support the more general hypotheses underlying this work that infections with viruses that use enzymes as their cell entry receptors can elicit the production of antibodies with physiological effects, antibodies with either catalytic activities or antibodies capable of inhibiting the activity of enzymes.

It may be considered whether the ACE2-like catalytic activity observed in the research subjects could be expected to mediate clinically meaningful changes in physiology. Unfortunately, this would be a very difficult undertaking in a study of banked samples and would probably require detailed physiological measurements on individual research subjects, paired with biochemical analyses of the bioactive peptides, and might even then be difficult. Angiotensinogen production varies considerably over time, and then so would its cleavage products, angiotensin II, and the presumed product of antibodies with the ACE2-like catalytic activity, angiotensin 1-7^47^. Angiotensin 1-7 has a very short half-life in the circulation. Reports differ, but the half-life could as short as a few seconds^48–50^. Some investigators have tried to develop angiotensin 1-7 as a therapeutic, but have had difficulties due to the short half-life and have, as result, been developing versions of angiotensin 1-7 conjugated to carriers to extend its half-life^51^. Since small changes in the radius of a vessel can lead to large changes in pressure (proportional to 1/r^4^), even small differences in vasoconstriction could yield physiologically detectable effects. Although our data showed some variation, we did detect the hypothesized impairment in physiologic homeostatic response following the mild exercise challenge (FIG 5), suggesting that an aberrant source of vasodilatory angiotensin 1-7 produced by abzymes with ACE2-like activity could blunt the normal homeostatic mechanisms that mediate the response to an exercise challenge. Much more detailed physiologic studies will be needed to further understand the clinical consequences of the existence of antibodies with ACE2-like catalytic activity.

Our study had several strengths. We studied a well-defined cohort of research volunteers, who had clear, persistent pulmonary symptoms after convalescing from acute COVID-19. Additional objective clinical and laboratory information was available for these individuals, enabling us to conduct preliminary analyses to determine whether there any clinical features that correlated with the presence of ACE2-like abzymes. We used a well-characterized ACE2 peptide substrate cleavage activity kit, and available size exclusion columns, protein A/G bead absorption immunoglobulin depletion procedures, and standard, available immunoglobulin assays.

Our study also has several limitations. We studied moderate number of volunteers with a single subset of post-COVID-19 clinical complaints, restrictive lung disease^44^. A much larger study or series of studies, involving many more research subjects with a variety of distinct principal manifestations of post-acute COVID-19 symptoms, ideally clearly defined LC symptomatology and additional, multiple comprehensive assays for antibody-associated catalytic activity would be needed to firmly support the abzyme hypothesis, but such work would be outside the scope of this study and samples that were available to us at the time of the study. The limitation of the cohort to those subjects with lung disease may also have limited any relationships between the presence of ACE2-like abzyme activity and physiological variables, particularly cardiovascular variables, since those are influenced by pulmonary disease. The persistent pulmonary symptoms that define this cohort likely result from many causes, perhaps most likely direct damage to lung parenchyma by viral replication and the secondary host inflammatory response. The cohort was not limited to volunteers who met a strict LC definition, so while some individuals had symptoms that could be characterized as belonging to the spectrum of LC, others in the cohort did not have such symptoms. Additional studies of a cohort specifically comprised of people with LC would be needed to determine whether LC symptoms were specifically associated with abzymes that had one or another catalytic activity. Other factors could certainly mediate the clinical symptoms that trouble people with LC. It is likely that LC symptoms have more than one cause. Among those other causes could be SARS-CoV-2 elicited abzymes with catalytic activities other than ACE2-like catalytic activities, but we were not able to test for those other activities in this study due to sample quantity constraints.

Finally, to definitively establish that an antibody induced by SARS-COV-2 has catalytic activity with potentially clinically meaningful effects, it would be necessary to isolate and study the antibody in pure form, something also outside the scope of this study.

Nonetheless, our study shows that some people convalescing from COVID-19 convalescent make antibodies with catalytic activity, a useful next step in understanding the pathogenesis of COVID-19 and LC.

## Institutional Review Board Statement

The University of Virginia Institutional Review Board for Health Sciences Research (FWA #00006183) approved enrollment of all subjects and collection of specimens and related metadata (HSR #13166) and approval was obtained to work on the specimens (HSR #HSR200362)

## Informed Consent Statement

Written informed consent was obtained from all subjects in the study.

## Data Availability Statement

Data is presented in the manuscript. Raw data is available upon request.

## Acknowledgments and Funding

The study was supported by internal funding from the University of Virginia, including the Manning Fund for COVID-19 Research at UVA the Ivy Foundation, the Pendleton Laboratory Fund for Pediatric Infectious Disease Research, and grants from NIAID, NIH (R01 AI176515, R21 AI160334, R56 AI178669). We thank the research volunteers for generously agreeing to be research subjects in the study, and Deborah Murphy RN who assisted with enrollment of study participants).

## Conflicts of Interest

The authors declare no conflict of interest.

## Notes

### Competing Interest Statement

The authors have declared no competing interest.

### Author Declarations

Institutional Review Board Statement: The University of Virginia Institutional Review Board for Health Sciences Research (FWA #00006183) approved enrollment of all subjects and collection of specimens and related metadata (HSR #13166) and approval was obtained to work on the specimens (HSR #HSR200362) Informed Consent Statement: Written informed consent was obtained from all subjects in the study.

### Summary of Updates

We submitted the manuscript for review. Anonymous reviewers requested changes and modification, which are reflected in this revision.

## References

1. Organization WH. WHO COVID-19 dashboard. World Health Organization; 2024 [cited 2024 24 June]; https://data.who.int/dashboards/covid19/deaths.

2. Altmann DM, Whettlock EM, Liu S, Arachchillage DJ, Boyton RJ. The immunology of long COVID. Nature Reviews Immunology. 2023;23(10):618–34.

3. Perumal R, Shunmugam L, Naidoo K, Abdool Karim SS, Wilkins D, Garzino-Demo A, Brechot C, Parthasarathy S, Vahlne A, Nikolich JŽ. Long COVID: a review and proposed visualization of the complexity of long COVID. Frontiers in immunology. 2023;14:1117464.

4. Davis HE, McCorkell L, Vogel JM, Topol EJ. Long COVID: major findings, mechanisms and recommendations. Nature Reviews Microbiology. 2023;21(3):133–46.

5. Adjaye-Gbewonyo D, Vahratian A, Perrine CG, Bertolli J. Long COVID in Adults: United States, 2022. 26/2023.

6. Castanares-Zapatero D, Chalon P, Kohn L, Dauvrin M, Detollenaere J, Maertens de Noordhout C, Primus-de Jong C, Cleemput I, Van den Heede K. Pathophysiology and mechanism of long COVID: a comprehensive review. Ann Med. 2022 Dec;54(1):1473–87. PMC9132392

7. Marchi M, Grenzi P, Serafini V, Capoccia F, Rossi F, Marrino P, Pingani L, Galeazzi GM, Ferrari S. Psychiatric symptoms in Long-COVID patients: a systematic review. Frontiers in Psychiatry. 2023;14:1138389.

8. Thaweethai T, Jolley SE, Karlson EW, Levitan EB, Levy B, McComsey GA, McCorkell L, Nadkarni GN, Parthasarathy S, Singh U, Walker TA, Selvaggi CA, Shinnick DJ, Schulte CCM, Atchley-Challenner R, Horwitz LI, Foulkes AS, Authors RC, Consortium R. Development of a Definition of Postacute Sequelae of SARS-CoV-2 Infection. JAMA. 2023;329(22):1934–46.

9. Soriano JB, Murthy S, Marshall JC, Relan P, Diaz JV. A clinical case definition of post-COVID-19 condition by a Delphi consensus. Lancet Infect Dis. 2022 Apr;22(4):e102–e7. PMC8691845

10. Klein J, Wood J, Jaycox JR, Dhodapkar RM, Lu P, Gehlhausen JR, Tabachnikova A, Greene K, Tabacof L, Malik AA, Silva Monteiro V, Silva J, Kamath K, Zhang M, Dhal A, Ott IM, Valle G, Peña-Hernández M, Mao T, Bhattacharjee B, Takahashi T, Lucas C, Song E, McCarthy D, Breyman E, Tosto-Mancuso J, Dai Y, Perotti E, Akduman K, Tzeng TJ, Xu L, Geraghty AC, Monje M, Yildirim I, Shon J, Medzhitov R, Lutchmansingh D, Possick JD, Kaminski N, Omer SB, Krumholz HM, Guan L, Dela Cruz CS, van Dijk D, Ring AM, Putrino D, Iwasaki A. Distinguishing features of long COVID identified through immune profiling. Nature. 2023 Nov;623(7985):139–48. PMC10620090

11. Cervia-Hasler C, Brüningk SC, Hoch T, Fan B, Muzio G, Thompson RC, Ceglarek L, Meledin R, Westermann P, Emmenegger M, Taeschler P, Zurbuchen Y, Pons M, Menges D, Ballouz T, Cervia-Hasler S, Adamo S, Merad M, Charney AW, Puhan M, Brodin P, Nilsson J, Aguzzi A, Raeber ME, Messner CB, Beckmann ND, Borgwardt K, Boyman O. Persistent complement dysregulation with signs of thromboinflammation in active Long Covid. Science. 2024 Jan 19;383(6680):eadg7942.

12. Huang C, Wang Y, Li X, Ren L, Zhao J, Hu Y, Zhang L, Fan G, Xu J, Gu X, Cheng Z, Yu T, Xia J, Wei Y, Wu W, Xie X, Yin W, Li H, Liu M, Xiao Y, Gao H, Guo L, Xie J, Wang G, Jiang R, Gao Z, Jin Q, Wang J, Cao B. Clinical features of patients infected with 2019 novel coronavirus in Wuhan, China. Lancet. 2020 Feb 15;395(10223):497–506. PMC7159299

13. Liao D, Zhou F, Luo L, Xu M, Wang H, Xia J, Gao Y, Cai L, Wang Z, Yin P, Wang Y, Tang L, Deng J, Mei H, Hu Y. Haematological characteristics and risk factors in the classification and prognosis evaluation of COVID-19: a retrospective cohort study. Lancet Haematol. 2020 Sep;7(9):e671–e8. PMC7351397

14. Del Valle DM, Kim-Schulze S, Huang HH, Beckmann ND, Nirenberg S, Wang B, Lavin Y, Swartz TH, Madduri D, Stock A, Marron TU, Xie H, Patel M, Tuballes K, Van Oekelen O, Rahman A, Kovatch P, Aberg JA, Schadt E, Jagannath S, Mazumdar M, Charney AW, Firpo-Betancourt A, Mendu DR, Jhang J, Reich D, Sigel K, Cordon-Cardo C, Feldmann M, Parekh S, Merad M, Gnjatic S. An inflammatory cytokine signature predicts COVID-19 severity and survival. Nat Med. 2020 Oct;26(10):1636–43. PMC7869028

15. Yousaf AR, Cortese MM, Taylor AW, Broder KR, Oster ME, Wong JM, Guh AY, McCormick DW, Kamidani S, Schlaudecker EP, Edwards KM, Creech CB, Staat MA, Belay ED, Marquez P, Su JR, Salzman MB, Thompson D, Campbell AP. Reported cases of multisystem inflammatory syndrome in children aged 12-20 years in the USA who received a COVID-19 vaccine, December, 2020, through August, 2021: a surveillance investigation. Lancet Child Adolesc Health. 2022 Feb 22. PMC8864018

16. De Rose DU, Pugnaloni F, Calì M, Ronci S, Caoci S, Maddaloni C, Martini L, Santisi A, Dotta A, Auriti C. Multisystem Inflammatory Syndrome in Neonates Born to Mothers with SARS-CoV-2 Infection (MIS-N) and in Neonates and Infants Younger Than 6 Months with Acquired COVID-19 (MIS-C): A Systematic Review. Viruses. 2022;14(4):750.

17. Oudit GY, Wang K, Viveiros A, Kellner MJ, Penninger JM. Angiotensin-converting enzyme 2-at the heart of the COVID-19 pandemic. Cell. 2023 Mar 2;186(5):906–22. PMC9892333

18. Yan R, Zhang Y, Li Y, Xia L, Guo Y, Zhou Q. Structural basis for the recognition of SARS-CoV-2 by full-length human ACE2. Science. 2020;367(6485):1444-8.

19. Borkotoky S, Dey D, Hazarika Z. Interactions of angiotensin-converting enzyme-2 (ACE2) and SARS-CoV-2 spike receptor-binding domain (RBD): a structural perspective. Mol Biol Rep. 2023 Mar;50(3):2713–21. PMC9786537

20. Wrobel AG. Mechanism and evolution of human ACE2 binding by SARS-CoV-2 spike. Curr Opin Struct Biol. 2023 Aug;81:102619. PMC10183628

21. Lan J, Ge J, Yu J, Shan S, Zhou H, Fan S, Zhang Q, Shi X, Wang Q, Zhang L, Wang X. Structure of the SARS-CoV-2 spike receptor-binding domain bound to the ACE2 receptor. Nature. 2020 2020/05/01;581(7807):215–20.

22. Wang Q, Zhang Y, Wu L, Niu S, Song C, Zhang Z, Lu G, Qiao C, Hu Y, Yuen K-Y, Wang Q, Zhou H, Yan J, Qi J. Structural and Functional Basis of SARS-CoV-2 Entry by Using Human ACE2. Cell. 2020;181(4):894–904.e9.

23. Khurana V, Goswami B. Angiotensin converting enzyme (ACE). Clin Chim Acta. 2022 Jan 1;524:113–22.

24. Turner AJ, Nalivaeva NN. Angiotensin-converting enzyme 2 (ACE2): Two decades of revelations and re-evaluation. Peptides. 2022 May;151:170766. PMC8830188

25. Tramontano A, Janda KD, Lerner RA. Catalytic antibodies. Science. 1986 Dec 19;234(4783):1566-70.

26. Pollack SJ, Jacobs JW, Schultz PG. Selective chemical catalysis by an antibody. Science. 1986 Dec 19;234(4783):1570-3.

27. Avalle B, Mistro D, Thomas D, Friboulet A. Polyclonal catalytic anti-idiotypic antibodies with a beta-lactamase activity. Ann N Y Acad Sci. 1996 Oct 12;799:172–5.

28. Friboulet A, Izadyar L, Avalle B, Roseto A, Thomas D. Abzyme generation using an anti-idiotypic antibody as the “internal image” of an enzyme active site. Appl Biochem Biotechnol. 1994 May-Jun;47(2-3):229–37; discussion 37-9.

29. Izadyar L, Friboulet A, Remy MH, Roseto A, Thomas D. Monoclonal anti-idiotypic antibodies as functional internal images of enzyme active sites: production of a catalytic antibody with a cholinesterase activity. Proc Natl Acad Sci U S A. 1993 Oct 1;90(19):8876–80. PMC47463

30. Friboulet A, Izadyar L, Avalle B, Roseto A, Thomas D. Antiidiotypic antibodies as functional internal images of enzyme-active sites. Ann N Y Acad Sci. 1995 Mar 31;750:265–70.

31. Ponomarenko NA, Pillet D, Paon M, Vorobiev, II, Smirnov IV, Adenier H, Avalle B, Kolesnikov AV, Kozyr AV, Thomas D, Gabibov AG, Friboulet A. Anti-idiotypic antibody mimics proteolytic function of parent antigen. Biochemistry. 2007 Dec 18;46(50):14598–609.

32. Hilvert D. Critical analysis of antibody catalysis. Annu Rev Biochem. 2000;69:751–93.

33. Bar-Even A, Noor E, Savir Y, Liebermeister W, Davidi D, Tawfik DS, Milo R. The moderately efficient enzyme: evolutionary and physicochemical trends shaping enzyme parameters. Biochemistry. 2011 May 31;50(21):4402–10.

34. Donoghue M, Hsieh F, Baronas E, Godbout K, Gosselin M, Stagliano N, Donovan M, Woolf B, Robison K, Jeyaseelan R, Breitbart RE, Acton S. A novel angiotensin-converting enzyme-related carboxypeptidase (ACE2) converts angiotensin I to angiotensin 1-9. Circ Res. 2000 Sep 1;87(5):E1–9.

35. Shuster AM, Gololobov GV, Kvashuk OA, Bogomolova AE, Smirnov IV, Gabibov AG. DNA hydrolyzing autoantibodies. Science. 1992 May 1;256(5057):665-7.

36. Paul S, Li L, Kalaga R, Wilkins-Stevens P, Stevens FJ, Solomon A. Natural catalytic antibodies: peptide-hydrolyzing activities of Bence Jones proteins and VL fragment. J Biol Chem. 1995 Jun 23;270(25):15257–61.

37. Paul S, Volle DJ, Beach CM, Johnson DR, Powell MJ, Massey RJ. Catalytic hydrolysis of vasoactive intestinal peptide by human autoantibody. Science. 1989 Jun 9;244(4909):1158-62.

38. Ponomarenko NA, Durova OM, Vorobiev, II, Belogurov AA, Jr., Kurkova IN, Petrenko AG, Telegin GB, Suchkov SV, Kiselev SL, Lagarkova MA, Govorun VM, Serebryakova MV, Avalle B, Tornatore P, Karavanov A, Morse HC, 3rd, Thomas D, Friboulet A, Gabibov AG. Autoantibodies to myelin basic protein catalyze site-specific degradation of their antigen. Proc Natl Acad Sci U S A. 2006 Jan 10;103(2):281–6. PMC1324791

39. Thiagarajan P, Dannenbring R, Matsuura K, Tramontano A, Gololobov G, Paul S. Monoclonal antibody light chain with prothrombinase activity. Biochemistry. 2000 May 30;39(21):6459–65.

40. Wootla B, Dasgupta S, Dimitrov JD, Bayry J, Lévesque H, Borg JY, Borel-Derlon A, Rao DN, Friboulet A, Kaveri SV, Lacroix-Desmazes S. Factor VIII hydrolysis mediated by anti-factor VIII autoantibodies in acquired hemophilia. J Immunol. 2008 Jun 1;180(11):7714–20.

41. Lacroix-Desmazes S, Moreau A, Sooryanarayana, Bonnemain C, Stieltjes N, Pashov A, Sultan Y, Hoebeke J, Kazatchkine MD, Kaveri SV. Catalytic activity of antibodies against factor VIII in patients with hemophilia A. Nat Med. 1999 Sep;5(9):1044–7.

42. Ponomarenko NA, Vorobiev, II, Alexandrova ES, Reshetnyak AV, Telegin GB, Khaidukov SV, Avalle B, Karavanov A, Morse HC, 3rd, Thomas D, Friboulet A, Gabibov AG. Induction of a protein-targeted catalytic response in autoimmune prone mice: antibody-mediated cleavage of HIV-1 glycoprotein GP120. Biochemistry. 2006 Jan 10;45(1):324–30.

43. Song Y, Myers R, Mehl F, Murphy L, Brooks B, Wilson JM, Kadl A, Woodfolk J, Zeichner SL. ACE-2-like enzymatic activity is associated with immunoglobulin in COVID-19 patients. mBio. 2024 Apr 10;15(4):e0054124. PMC11005375

44. Canderan G, Muehling LM, Kadl A, Ladd S, Bonham C, Cross CE, Lima SM, Yin X, Sturek JM, Wilson JM, Keshavarz B, Enfield KB, Ramani C, Bryant N, Murphy DD, Cheon IS, Solga M, Pramoonjago P, McNamara CA, Sun J, Utz PJ, Dolatshahi S, Irish JM, Woodfolk JA. Distinct type 1 immune networks underlie the severity of restrictive lung disease after COVID-19. Nat Immunol. 2025 Apr;26(4):595–606.

45. Keshavarz B, Wiencek JR, Workman LJ, Straesser MD, Muehling LM, Canderan G, Drago F, Bonham CA, Sturek JM, Ramani C, McNamara CA, Woodfolk JA, Kadl A, Platts-Mills TAE, Wilson JM. Quantitative Measurement of IgG to Severe Acute Respiratory Syndrome Coronavirus-2 Proteins Using ImmunoCAP. Int Arch Allergy Immunol. 2021;182(5):417–24. PMC8018212

46. Geanes ES, McLennan R, LeMaster C, Bradley T. Autoantibodies to ACE2 and immune molecules are associated with COVID-19 disease severity. Communications Medicine. 2024 2024/03/15;4(1):47.

47. Paul M, Poyan Mehr A, Kreutz R. Physiology of local renin-angiotensin systems. Physiological reviews. 2006;86(3):747–803.

48. Jiang F, Yang J, Zhang Y, Dong M, Wang S, Zhang Q, Liu FF, Zhang K, Zhang C. Angiotensin-converting enzyme 2 and angiotensin 1–7: novel therapeutic targets. Nature Reviews Cardiology. 2014 2014/07/01;11(7):413–26.

49. Touyz RM, Montezano AC. Angiotensin-(1–7) and Vascular Function. Hypertension. 2018 2018/01/01;71(1):68–9.

50. McKinney Clare A, Fattah C, Loughrey Christopher M, Milligan G, Nicklin Stuart A. Angiotensin-(1–7) and angiotensin-(1–9): function in cardiac and vascular remodelling. Clinical Science. 2014;126(12):815–27.

51. Khajeh Pour S, Ranjit A, Summerill EL, Aghazadeh-Habashi A. Anti-Inflammatory Effects of Ang-(1-7) Bone-Targeting Conjugate in an Adjuvant-Induced Arthritis Rat Model. Pharmaceuticals (Basel). 2022 Sep 17;15(9). PMC9502795

